# Perfusion imaging heterogeneity during NO inhalation distinguishes pulmonary arterial hypertension (PAH) from healthy subjects and has potential as an imaging biomarker

**DOI:** 10.1101/2022.08.16.22278842

**Authors:** Tilo Winkler, Puja Kohli, Vanessa J Kelly, Ekaterina G. Kehl, Alison S. Witkin, Josanna Rodriguez-Lopez, Kathryn A. Hibbert, Mamary Kone, David M. Systrom, Aaron B. Waxman, Jose G. Venegas, Richard Channick, R. Scott Harris

**Affiliations:** Department of Anesthesia, Critical Care and Pain Medicine, Massachusetts General Hospital and Harvard Medical School, Boston, Massachusetts, USA; Division of Pulmonary and Critical Care Medicine, Department of Medicine, Massachusetts General Hospital and Harvard Medical School, Boston, Massachusetts, USA; Division of Pulmonary and Critical Care Medicine, Department of Medicine, Brigham and Women’s Hospital and Harvard Medical School, Boston, Massachusetts, USA

**Keywords:** Pulmonary Circulation, Vascular Physiology, Functional Imaging, Positron Emission Tomography, Perfusion distribution, Ventilation/Perfusion distribution, Inhaled nitric oxide

## Abstract

**Background:** Without aggressive treatment, pulmonary arterial hypertension (PAH) has a 5-year mortality of approximately 40%. A patient’s response to vasodilators at diagnosis impacts the therapeutic options and prognosis. We hypothesized that analyzing perfusion images acquired before and during vasodilation could identify characteristic differences between PAH and control subjects.

**Methods:** We studied 5 controls and 4 subjects with PAH using HRCT and ^13^NN PET imaging of pulmonary perfusion and ventilation. The total spatial heterogeneity of perfusion (CV^2^_Qtotal_) and its components in the vertical (CV^2^_Qvgrad_) and cranio-caudal (CV^2^_Qzgrad_) directions, and the residual heterogeneity (CV^2^_Qr_), were assessed at baseline and while breathing oxygen and nitric oxide (O_2_+iNO). The length scale spectrum of CV^2^_Qr_ was determined from 10 to 110 mm, and the response of regional perfusion to O_2_+iNO was calculated as the mean of absolute differences. Vertical gradients in perfusion (Q_vgrad_) were derived from perfusion images, and ventilation-perfusion distributions from images of ^13^NN washout kinetics.

**Results:** O_2_+iNO significantly enhanced perfusion distribution differences between PAH and controls, allowing differentiation between PAH subjects from controls. During O_2_+iNO, CV^2^_Qvgrad_ was significantly higher in controls than in PAH (0.08 (0.055-0.10) vs. 6.7 × 10^−3^ (2×10^−4^-0.02), p<0.001) with a considerable gap between groups. Q_vgrad_ and CV^2^_Qtotal_ showed smaller differences: –7.3 vs. -2.5, p = 0.002, and 0.12 vs. 0.06, p = 0.01. CV^2^_Qvgrad_ had the largest effect size among the primary parameters during O_2_+iNO. CV^2^_Qr_, and its length scale spectrum were similar in PAH and controls. Ventilation-perfusion distributions showed a trend towards a difference between PAH and controls at baseline, but it was not statistically significant.

**Conclusions:** Perfusion imaging during O2+iNO showed a significant difference in the heterogeneity associated with the vertical gradient in perfusion, distinguishing in this small cohort study PAH subjects from controls.

**Trial registration:** Not applicable

## INTRODUCTION

Pulmonary arterial hypertension (PAH) is a progressive disease with a 5-year mortality of approximately 40% [1]. Wide variations in the effectiveness of vasodilator treatment is a major challenge to improving outcomes [2]. Treatment guidelines define a positive response to vasodilator challenge as a reduction of ≥ 10 mmHg in the mean pulmonary arterial pressure (mPAP) to reach an absolute value of mPAP ≤ 40 mmHg with an increased or unchanged cardiac output [3], however, only about 10% of patients with idiopathic PAH meet these clinical criteria for a response to vasodilators [3] that are associated with greater treatment options and better survival [2,3]. Interestingly, it has been shown that the magnitude of responses in pulmonary vascular resistance (PVR) or mean PAP to inhaled nitric oxide with oxygen predicts long-term survival even among patient that are non-responders [4]. Overall, however, we have little insight into the pulmonary vascular function that is associated with increased risk.

In contrast to global parameters such as mPAP or pulmonary vascular resistance (PVR), imaging of pulmonary perfusion provides insights into regional changes associated with the disease process and its heterogeneity affecting the perfusion distribution inside the lungs. However, it is unclear if pulmonary perfusion imaging can detect substantial differences between PAH and healthy controls or assess regional changes in response to acute vasodilators such as inhaled nitric oxide and oxygen. In healthy lungs, there is a vertical gradient in perfusion [5,6] including reduced lung inflation of dependent lung regions due to gravity [7]. Additionally, there is heterogeneity in perfusion within isogravitational slices that might be related to the fractal nature of the distribution of alveoli and the structure of the pulmonary vascular tree [8–10]. In addition, super-imposed disease processes can lead to regional changes in lung structure or function with characteristic sizes reflected in the length-scale spectrum of perfusion heterogeneity. It is unclear if underlying vascular structural changes in PAH, such as plexiform lesions [11], increase the regional heterogeneity of perfusion. We previously showed that length-scale analysis of pulmonary perfusion images could detect exercise PAH at rest [12]. Other imaging studies found associations between PAH and changes in the redistribution of pulmonary perfusion in response to changes in body position [13,14], regional pulmonary blood volume [15–17], pulmonary impedance associated with perfusion [18], and the vertical gradient in pulmonary perfusion [19]. Though early detection of PAH could improve the survival of patients [20], no pulmonary imaging parameter has been identified yet to allow a precise differentiation between PAH patients and healthy individuals.

Here, we used positron-emission tomography and high-resolution computer tomography (PET-CT) imaging characterize the spatial distributions of pulmonary perfusion and ventilation in patients with PAH. We compared the patterns of perfusion while breathing air and during inhalation of nitric oxide (iNO) and both oxygen (O_2_) and inhaled nitric oxide (O_2_+iNO) to differentiate active effects of vasoconstriction [21], anatomical structure of the vascular tree and the vertical gradient in hydrostatic pressure due to gravity. We hypothesized that differences in pulmonary perfusion images acquired at baseline and during vasodilation could identify characteristic perfusion patterns between controls and PAH subjects, even in PAH subjects deemed “non-responders” to acute vasodilation during cardiac catheterization.

## METHODS

### Subjects

The study was approved by the Institutional Review Board of the Massachusetts General Hospital. Informed consent was obtained from each subject before the study. We previously published an imaging study of exercise PAH [12], using the same control group as in the present study. However, there is no overlap in primary parameters and findings between both studies. Subjects enrolled with PAH at the time of recruitment, as defined by the World Health Organization Group 1 classification, were required to have a right heart catheterization (RHC) within 12 months of study entry demonstrating mPAP >25 mmHg and pulmonary capillary wedge pressure (PCWP) < 15 mm Hg.

Spirometry data were collected for subjects with PAH. Trends in cardiac output were non-invasively recorded in all subjects using impedance cardiography (ICON, Cardiotronic, LaJolla, CA). Due to the known limitations of impedance cardiography, a trend rather than an absolute change in cardiac output, was monitored. For assessment of within-subject changes in perfusion and perfusion heterogeneity compared to baseline and for the comparison of regional perfusion between PAH and controls, we used values of mean-normalized regional perfusion obtained directly from imaging data. Pulse oximetry and heart rate were continuously monitored from a fingertip pulse oximeter.

PAH subjects receiving vasodilators as part of their treatment were maintained on those therapies during the day of study.

### Image Acquisition

A catheter was placed in an antecubital vein for radioisotope injection. Subjects were positioned supine and with arms abducted on the table of the PET-CT scanner (Biograph 64, Siemens Healthcare, Malvern PA, USA). Relative lung volume was continuously measured using impedance plethysmography (SomnoStar PT, SensorMedics Corp., Yorba Linda, CA, USA). After calibration of the plethysmograph, mean lung volume (MLV) was determined during 30 seconds of steady tidal breathing prior to imaging. Image acquisition started a ‘topogram’ CT scan to determine the field of view for PET and HRCT scans. In preparation for an HRCT scan, the subject was asked to inhale to total lung capacity (TLC), followed by slow exhalation, and to hold the breath when the impedance plethysmography signal showed that MLV was reached. During the breath-hold the HRCT scan was acquired. Following this, a dynamic ^13^NN-saline PET scan was acquired starting with an equivalent breath-hold at MLV. Simultaneously with the beginning of the breath-hold we injected a bolus of ^13^NN dissolved in saline (25mL) and started the acquisition of the dynamic PET scan. After 30 seconds of breath-hold, we asked the subject to resume breathing while the PET scan acquisition continued. Total acquisition time of the PET scan was 6 minutes and 40 seconds. During this first PET-CT imaging sequence the subject was breathing air. After it was completed, subjects were administered a gas mixture (iNO, iNOMax Delivery System) of balance gas oxygen with 30 ppm inhaled nitric oxide (O_2_+iNO) through a one-way valve with a mouthpiece and nose clip. After at least 5 minutes of breathing O_2_+iNO, we repeated the PET-CT imaging sequence while the subject continued to breath the O_2_+iNO gas mixture. Given the sample size and the desire to prevent carry over effects from the inhaled nitric oxide, all subjects underwent baseline imaging followed by imaging with iNO without randomization of order of imaging.

HRCT scans (helical mode with 64 slices per rotation, 120 kVp, and 80 mAs) were reconstructed with filtered back-projection (convolution kernel B31f, 0.5 mm slice increment, voxel size: 0.7324 × 0.7324 × 0.5 mm^3^). The PET field of view included on average 75% of the CT volume of the thorax. Dynamic PET scans were reconstructed with filtered back-projection and attenuation corrections using the HRCT scan. The dynamic PET scans yielded a stack of 19 images (8×5s, 3×10s, 7×30s, 60s, 2×30s), each image a 3D matrix of 128×128×81 voxels (voxel size 5.3456 × 5.3456 × 2.0250 mm^3^). The PET scans have a full-width at half maximum (FWHM) resolution of 6 mm (Biograph 64, Siemens Healthcare, Malvern PA, USA) so that the effective resolution with a contrast recovery coefficient of 80% at two times FWHM is approximately 12 mm. PET images were filtered before image analysis using a moving average filter of five slices (∼10mm) in the axial direction and a filter with 10 mm diameter within slices, which is comparable to the effective resolution of the scans.

### Image Analysis

Image segmentation software Apollo (VIDA Diagnostics, IA, USA) was used to identify the lung region of interest (ROI) in the HRCT scans. Using custom software tools written in Matlab (Mathworks, Natick, MA), the lung ROIs were converted to ROI masks for PET scan analysis. Additionally, HRCT scans were converted into gas fraction (F_gas_) images using the equation:

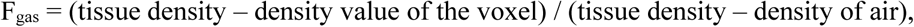

where tissue density = 0 Hounsfield units (HU) and density of gas = -1000 HU. Tissue fraction (F_tis_) images were calculated using F_tis_ = 1 – F_gas_.

Perfusion and ventilation images at baseline and after O_2_+iNO were generated from ^13^NN-saline PET scans. The analysis of dynamic PET scans of ^13^NN kinetics following a bolus injection have been described previously [22,23]. Briefly, nitrogen gas has a very low solubility, and intravenously injected ^13^NN, dissolved in saline, diffuses during its passage through the pulmonary capillaries rapidly out of the blood into the alveolar gas space, where it accumulates during the breath hold reaching a plateau value that is directly proportional to the local fraction of perfusion. Thus, the voxel values of a PET image of ^13^NN activity during the plateau, divided by the mean activity within the lung represents those of a mean-normalized perfusion distribution, allowing straightforward comparisons among images taken at different conditions. After the breath hold, the subject resumes tidal breathing resulting in the washout of the ^13^NN by ventilation with the tracer kinetics during this period yielding an estimate of specific ventilation 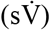 defined as: 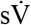 = alveolar ventilation 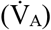 alveolar gas volume.

Assessment of regional ventilation and 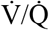 ratio using ^13^NN tracer kinetics modelling was performed as previously described [23]. Briefly, the tracer kinetics curves of each voxel were analysed to determine 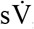, the perfusion associated with the 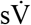 kinetics, and 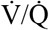 ratio. Four models were used for quantification of tracer washout kinetics: 1) single-exponential washout describing homogenous voxels, 2) complete gas-trapping for voxels with no change in activity during washout, 3) a double-exponential washout model describing heterogenous voxels with two compartments that have different 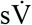, and 4) a partial gas trapping model for heterogeneous voxels with one washout and one gas trapping compartment. The perfusion associated with the 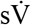 of a voxel or compartment was proportional to the initial activity of the tracer kinetics. After fitting all four models for each voxel, we used Akaike Information Criterion (AIC) (32) to select for each individual voxel the optimal model yielding 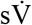 and perfusion of the voxel or sub-voxel compartments. 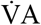 A was calculated as 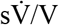, where V = Fgas * voxel volume. For voxels with two compartments, the gas volume of each compartment was assumed to be proportional to the fraction of blood flow in this compartment relative to the whole voxel: V_i_ = Fgas * voxel volume * Q_i_ / Q_voxel_, were i is compartment 1 or 2, and Q_voxel_ = Q_1_ + Q_2_. Mean normalized 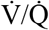 distributions were computed from the 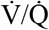 ratios of the individual voxels and sub-voxel compartments using 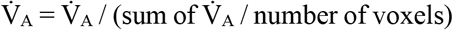 and Q = Q / (sum of Q / number of voxels). 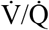 plots were constructed by binning of the typically 20,000-50,000 individual voxel and sub-voxel 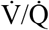 values using 100 bins over the 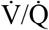 range from 0.001 to 100. Note that the voxels with two-compartment washout kinetics were included as independent compartments [24,25]. The 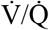 dispersions of perfusion SD Q 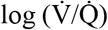 and ventilation SD V 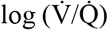were calculated as the standard deviations of the perfusion-weighted and ventilation-weighted natural logarithm (Peter Wagner, personal communication, July 11, 2020) of the 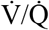 distribution, respectively [26,27]. Note that other authors have used the term log SD Q, but we wanted to avoid a misinterpretation as log(SD(Q)).

### Perfusion Heterogeneity and Length Scale Spectrum Analysis

Heterogeneity of perfusion was determined as the square of the coefficient of variation (CV^2^ = (standard deviation/mean)^2^) using a previously developed and validated method for assessment of noiseless CV^2^ of PET scans based on theoretical properties of measurement errors and empirical assessment of imaging noise [28]. In short, random errors in the individual voxels of PET images, referred to as imaging noise, are superimposed to the true voxel values (signal) of the PET image and uncorrelated with the imaging signal so that the heterogeneity of an image including noise (CV^2^_Qimage_) is the sum of the variance of the true signal (perfusion) and the variance of the imaging noise CV^2^_Qimage_ = CV^2^_Qtotal_ + CV^2^_noise_ [28]. We estimated CV^2^_noise_ using multiple PET images during the tracer plateau at the end of the breath-hold period and calculated the total spatial heterogeneity of ^13^NN images of relative perfusion, excluding the imaging noise, as CV^2^_Qtotal_ = CV^2^_Qimage_ – CV^2^_noise_. In the supine position, gravitational forces typically cause a gradient in perfusion in the dorsoventral direction with higher perfusion in the dependent (dorsal) regions than in the non-dependent (ventral) regions, which can be a substantial component of CV^2^_Qtotal_. We separated the total spatial heterogeneity of perfusion into components caused by systematic gradients in the vertical direction (CV^2^_Qvgrad_), the axial (craniocaudal) direction (CV^2^_Qzgrad_), and the remaining, or residual heterogeneity (CV^2^_Qr_). Thus, CV^2^_Qtotal_ = CV^2^_Qvgrad_ + CV^2^_zgrad_ + CV^2^_Qr_. The value of CV^2^_Qr_ was further split into its components due to spatial variations in perfusion across the length scale spectrum [23,25]. Such a length scale spectrum quantifies the components of heterogeneity originating from spatial patterns in regional perfusion measured at different image resolutions.

For each condition, the length scale spectrum of perfusion was quantified as follows: First, the vertical and the axial gradients (z direction) in perfusion (CV^2^_Qvgrad_ and CV^2^_Qzgrad_) were determined. Then the gradients were subtracted voxel-by-voxel from the 3D perfusion scan, and each of the resulting transverse slices of the scan were low-pass filtered at 10, 30, 50, 70, 90 and 110 mm with correction for edge effects [25], CV^2^ was calculated for each filter size, and the contribution of each length scale ranges were calculated as differences between successive filter sizes, e.g. CV^2^_Q10-30_ = CV^2^(10mm) – CV^2^(30mm). Overall, small length scales of a spectrum starting at 10 mm correspond to variation among volumes of approximately 1 mL, which corresponds to about 5 acinar units; whereas large length scales of the spatial spectrum correspond to heterogeneity due to variation in structural, functional and disease processes among larger regions such as segments or subsegments.

### Voxel Mapping and Changes in Regional Perfusion

For the assessment of changes in perfusion at the voxel level, we performed elastic image registration using the Advanced Normalization Tools (ANTs, free and open-source software, https://stnava.github.io/ANTs/) to correct for potential tissue displacement between the two perfusion images. First, we identified the transformation field that mapped the HRCT scan taken at MLV while breathing O_2_+iNO to the corresponding HRCT scan taken at MLV while breathing air. The same method has been successfully used before to determine the regional volumetric deformation and regional lung strain during mechanical ventilation [29,30]. In principle, this elastic registration method uses the density information of the HRCT scans for the identification of the transformation field T(x) describing the change in tissue location from one HRCT scan to the other. Image acquisition for both CT and PET scans at MLV allows for the application of the transformation field T(x) to the O_2_+iNO PET scan to map its voxels to the corresponding voxels of the air PET scan correcting differences in tissue location. This correction is essential for an assessment of changes between PET scans at the voxel level. Images representing the relative changes in regional perfusion between baseline and O_2_+iNO were calculated as voxel-by-voxel differences between the mean-normalized perfusion at air and the mean-normalized perfusion at O_2_+iNO.

Perfusion-height maps were generated as two-dimensional (2D) histograms of the number of voxels in relation to relative perfusion and relative height from the most dorsal point of the lung mask and the most ventral point. Difference maps of the perfusion-height distributions between air and O_2_+iNO were calculated as the difference between the perfusion-height distribution at O_2_+iNO and the distribution at air. Values in the difference map are visualized with a grey scale: no change equals 50% grey, decrease in perfusion is lighter, and an increase is darker.

Mean absolute changes in regional perfusion between air and O_2_+iNO were calculated at the voxel level as mean absolute deviations. Contributions of imaging noise [28] to the mean deviations were removed using the statistical properties of the difference image. Specifically, total variance of the difference image of perfusion between air and O_2_+iNO includes the sum of the variances of imaging noise in the two original images. The variance of the imaging PET noise, which has a normal distribution, was determined, using a previously described method for noise assessment using multiple time frames of a PET scan [28], and subtracted from the total variance of the difference image. The mean absolute deviation was then calculated using the relationship to the variance for a normal distribution as square root of two over pi times the variance. In order to rule out that imaging noise could explain the variations in the difference image, we tested in each individual study if the overall variance of the difference image was higher than the upper limit of the 99.9% confidence interval of the variance from noise.

### Statistical Analysis

Our primary outcomes of interest were the noise corrected values of total spatial heterogeneity of perfusion (CV^2^_Qtotal_) and its components, including the heterogeneity generated by the vertical gradient in perfusion (CV^2^_Qvgrad_) and the residual perfusion heterogeneity (CV^2^_Qr_) and its length scale components. In secondary analyses, we assessed various regional perfusion and ventilation metrics. We performed two-sided T-tests to evaluate whether there was a difference in means of each metric between study groups and for baseline vs. O_2_+iNO within study groups. Additionally, we obtained Cohen’s d effect size estimates for the primary parameters. All analyses were performed using R Statistical Software (v4.1.3)[31]. Effect sizes were estimated using the effsize R package (v0.8.1)[32]. Statistical significance was set at p < 0.05. Data are presented as mean (range) unless otherwise stated.

## RESULTS

### Study Participants

Nine subjects were studied: 5 controls, and 4 subjects with PAH (**Table 1**). Subjects were free of infections or rapid progression of their disease for at least one month prior to the study and did not have any other known cardiac or pulmonary disease. No subjects required oxygen therapy at the time of imaging. All controls were non-smokers with subjectively normal exercise tolerance and normal pulmonary function tests. All PAH subjects were initially found to have normal pulmonary function testing and normal chest imaging but two of them were subsequently found to have mildly abnormal spirometry on a repeat testing after imaging. PAH subjects had elevated resting mPAP (mean 43; range 33-63 mmHg) with normal PCWP (mean 11; range 7-18 mmHg). Two PAH subjects were “non-responders” as demonstrated by absence of significant reduction in mean pulmonary artery pressure and increased cardiac output after administration of a short-acting vasodilator, one was a responder, and one did not have a standardized test. Cardiac outputs were lower while breathing O_2_+iNO for both groups. However, there were no significant differences in cardiac output trends between groups at rest or while breathing O_2_+iNO. All PAH subjects were on combination therapy with a PDE5 inhibitor and endothelin receptor antagonist, and two PAH subjects were also on an inhaled prostacyclin or a prostacyclin receptor agonist.

**Table 1.** Baseline characteristics.

| **Parameter** | **Control**<br>N=5 | **PAH**<br>N=4 | **p-value** |
| --- | --- | --- | --- |
| **Age (years)** | 41.2 (22-60) | 47.3 (22-64) | 0.64 |
| **Gender, Female** | 1 | 3 | 0.14 |
| **Race, White** | 5 | 3 | 0.39 |
| **Height (inches)** | 67.3 (63-71.5) | 67.2 (66-68.5) | 0.91 |
| **Weight (pounds)** | 189.6 (160-296.7) | 170.5 (124-264) | 0.67 |
| **Years of symptoms** | NA | 8.4 (7-10) |  |
| **FEV1 (%predicted)** | NA | 82.9 (66-95) |  |
| **FVC (% predicted)** | NA | 85.6 (64-110) |  |
| **FEV1/FVC** | NA | 80 (63-86) |  |
| **mPAP, rest (mmHg)** | NA | 41.6 (33-63) |  |
| **PVR, rest (dyn·s/cm<sup>5</sup>)** | NA | 442 (322-548) |  |
| **PCWP, rest (mmHg)** | NA | 10 (7-16) |  |
| **6MWD (feet)** | NA | 1519.8 (1253-2198) |  |

Continuous variables are represented as mean (min-max). Categorical variables are represented as counts.
Definitions of abbreviations: PAH = primary pulmonary arterial hypertension; FEV<sub>1</sub> = forced expiratory volume in first second; FVC= forced vital capacity; mPAP = mean pulmonary arterial pressure; PVR = pulmonary vascular resistance; PCWP = pulmonary capillary wedge pressure; 6MWD = six-minute walk distance.

### Regional Perfusion Distribution: Effect of iNO and Oxygen

#### Total Spatial Heterogeneity in Perfusion

At baseline, the total spatial heterogeneity of perfusion (CV^2^_Qtotal_) for the PAH subjects was similar to that of the controls (p=0.51). However, while breathing O_2_+iNO, the PAH subjects had a lower CV^2^_Qtotal_ (2-fold, p=0.01) compared to control counterparts (Figure 1, **Table 2**).

**Table 2.** Vertical gradient and heterogeneity in perfusion.

| Parameter | Control |  | PAH |  |
| --- | --- | --- | --- | --- |
|  | Baseline | O<sub>2</sub>+iNO | Baseline | O<sub>2</sub>+iNO |
| **Q<sub>vgrad</sub> (10<sup>-2</sup> cm<sup>-1</sup>)** | -5.46 (-3.32, -7.01)*<sup>†</sup> | -7.37 (-6.43, -8.66)*<sup>‡</sup> | -3.13 (-2.33, -4.78)<sup>†</sup> | -2.51 (-0.039, -0.431)<sup>‡</sup> |
| **CV<sup>2</sup><sub>Qtotal</sub>** | 0.09 (0.06, 0.16) | 0.12 (0.08, 0.16)<sup>‡</sup> | 0.08 (0.07, 0.11) | 0.06 (0.04, 0.1)<sup>‡</sup> |
| **CV<sup>2</sup><sub>Qvgrad</sub>** | 0.04 (0.02, 0.08)<sup>†</sup> | 0.08 (0.055, 0.10)<sup>‡</sup> | 0.01 (0.0, 0.03)<sup>†</sup> | 0.01 (0.0002, 0.02)<sup>‡</sup> |
| **CV<sup>2</sup><sub>Qr</sub>** | 0.04 (0.02, 0.07) | 0.04 (0.02, 0.06) | 0.05 (0.04, 0.06) | 0.03 (0.02, 0.04) |

Continuous variables are represented as mean (min, max).
Definitions of abbreviations: PAH = primary pulmonary arterial hypertension; Q<sub>vgrad</sub> = vertical gradient in regional perfusion (slope); CV<sup>2</sup><sub>Qtotal</sub> = squared coefficient of variation in perfusion quantifying total spatial heterogeneity of perfusion (noise corrected); CV<sup>2</sup><sub>Qvgrad</sub> = squared coefficient of variation in perfusion quantifying heterogeneity generated by the dorsal-ventral gradient in perfusion; CV<sup>2</sup><sub>Qr</sub> = squared coefficient of variation in perfusion quantifying heterogeneity of the residual perfusion.
\* significant difference between measures at baseline and during administration of O<sub>2</sub>+iNO within a group
<sup>†</sup> significant difference between PAH subjects and controls at baseline
<sup>‡</sup> significant difference between PAH subjects and controls while breathing O<sub>2</sub>+iNO

**Figure 1:**
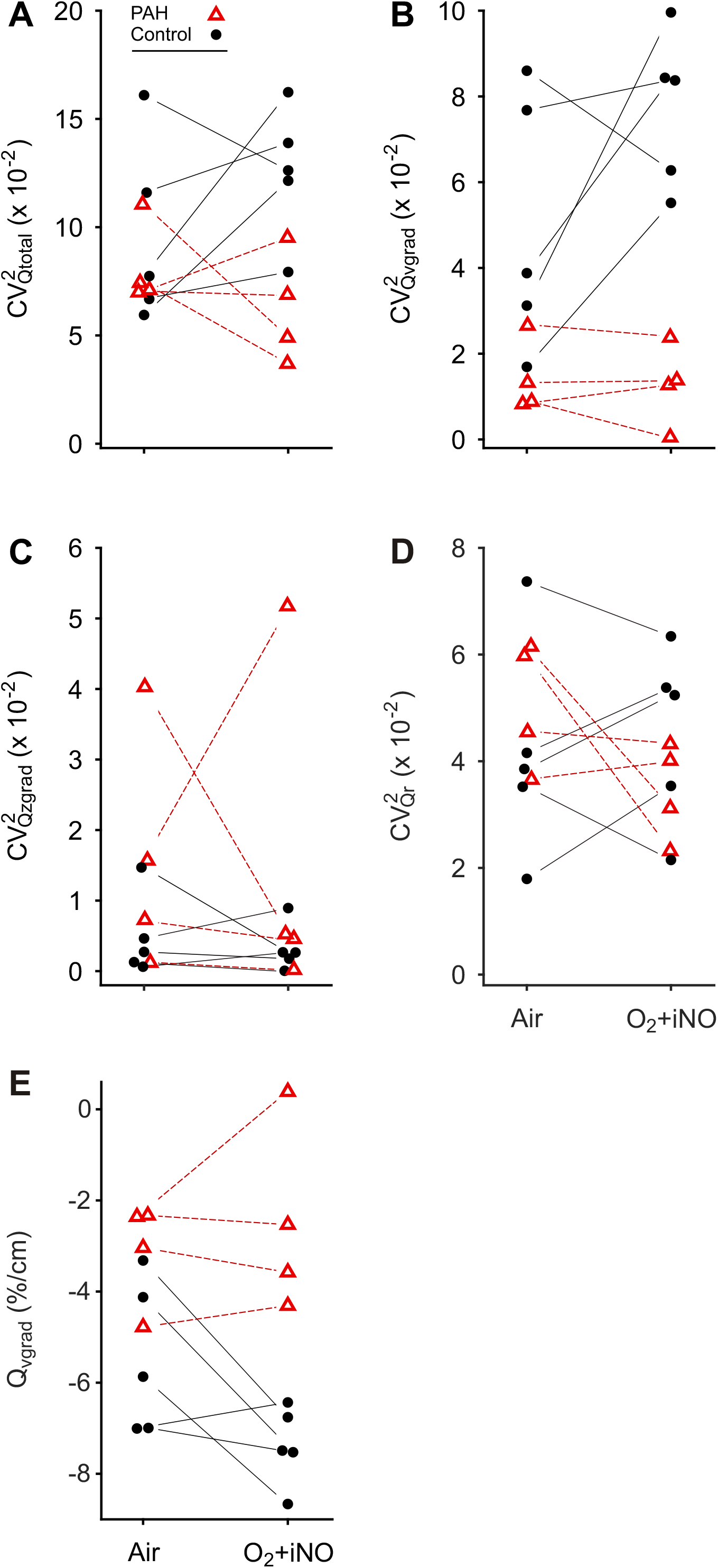
Perfusion heterogeneity profiling distinguished PAH subjects and control subjects. The total spatial heterogeneity of perfusion (CV^2^_Qtotal_)[28] (**A**) is comprised of the heterogeneity of the vertical (dorsoventral) gradient in perfusion (CV^2^_Qvgrad_) (**B**), the heterogeneity of the craniocaudal (z-axis) gradient in perfusion (CV^2^_Qzgrad_) (**C**), and the heterogeneity of the residual (or remaining) perfusion (CV^2^_Qr_) (**D**). PAH subjects demonstrated very low CV^2^_Qvgrad_ due to attenuated vertical gradients in perfusion (Q_vgrad_)(**E**) that were not responsive to O_2_+iNO, distinguishing this cohort from their control counterparts. Note that imaging noise is excluded from all CV^2^ parameters.

#### Vertical and Axial Gradients in perfusion and the associated heterogeneity

A vertical gradient in regional perfusion was observed in both groups, with higher perfusion in the dependent regions (dorsal) relative to the non-dependent (ventral) regions (Figure 1, **Table 2**). At baseline breathing air, the magnitude of the vertical gradient (Q_vgrad_) and the heterogeneity generated by that gradient (CV^2^_Qvgrad_) were significantly greater in the control group compared with the PAH group (p=0.04 and 0.05, respectively) (**Table 2**). During O_2_+iNO, these differences between groups were enhanced: CV^2^_Qvgrad_ was 6.1-fold and Q_vgrad_ 2.9-fold, greater in the controls compared to the PAH subjects (p<0.001 and p=0.013, respectively) (Figure 1, **Table 2**). Notably, during O_2_+iNO, both CV^2^_Qvgrad_ and Q_vgrad_ distinguished PAH subjects from controls with a considerable gap between groups compared to the overlap at baseline breathing air (Figure 1). Quantifying the effect size using Cohen’s d, CV^2^_Qvgrad_ during O_2_+iNO showed the largest estimate with a 95%-confidence interval not crossing zero, followed by Q_vgrad_ and CV^2^_Qtotal_: 4.30 (1.42, 7.17), –3.25 (–5.65, –0.84) and 2.23 (0.22, 4.25), respectively (Figure 2). In the individual subjects, the average response of CV^2^_Qvgrad_ to breathing O_2_+iNO was an increase of 0.0272 in the controls compared to a minimal decrease of -0.0017 in PAH subjects. Our study was not designed to assess differences between responders and non-responders in PAH. Nevertheless, we noted that the only confirmed responder among the PAH subjects showed an increase in CV^2^_Qvgrad_ during O_2_-iNO in the contrast to decreases in CV^2^_Qvgrad_ in the other three PAH subjects (Figure 1B). PAH subjects generally displayed low vertical gradients in perfusion at baseline and during O_2_+iNO without a significant change in the gradient (**Figures 3**). Also, the differences in the vertical gradient in perfusion between groups persisted after adjusting the regional blood flow by regional lung density determined from HRCT.

**Figure 2:**
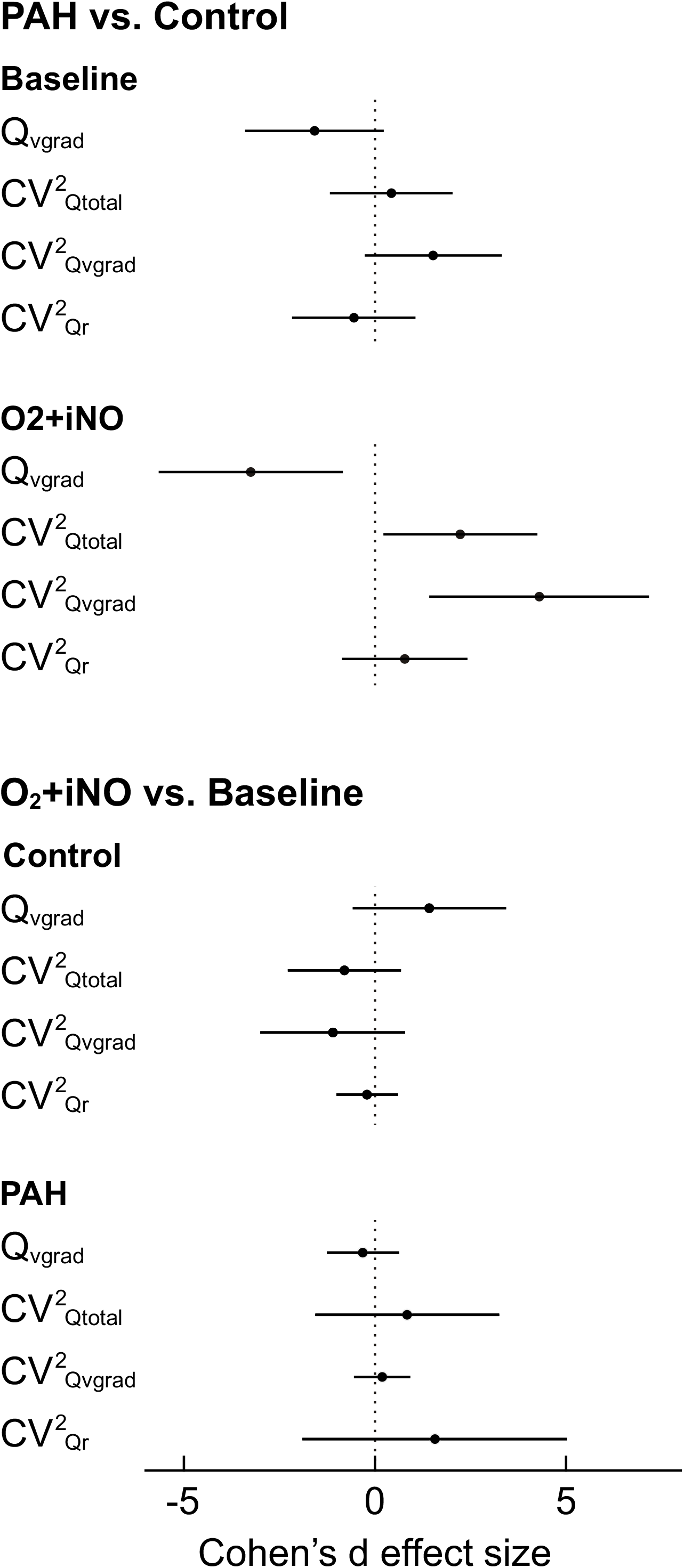
Cohen’s d effect size estimates and 95% confidence intervals of the primary parameters. The effect sizes for PAH vs. control were substantially larger for O_2_+iNO compared to baseline. CV^2^_Qvgrad_ had the most significant effect size and a 95% confidence interval not crossing zero, followed by Q_vgrad_ and CV^2^_Qtotal_. The relatively large effect sizes for O_2_+iNO vs. baseline in the control group are indicators of the magnitude of the response.

**Figure 3:**
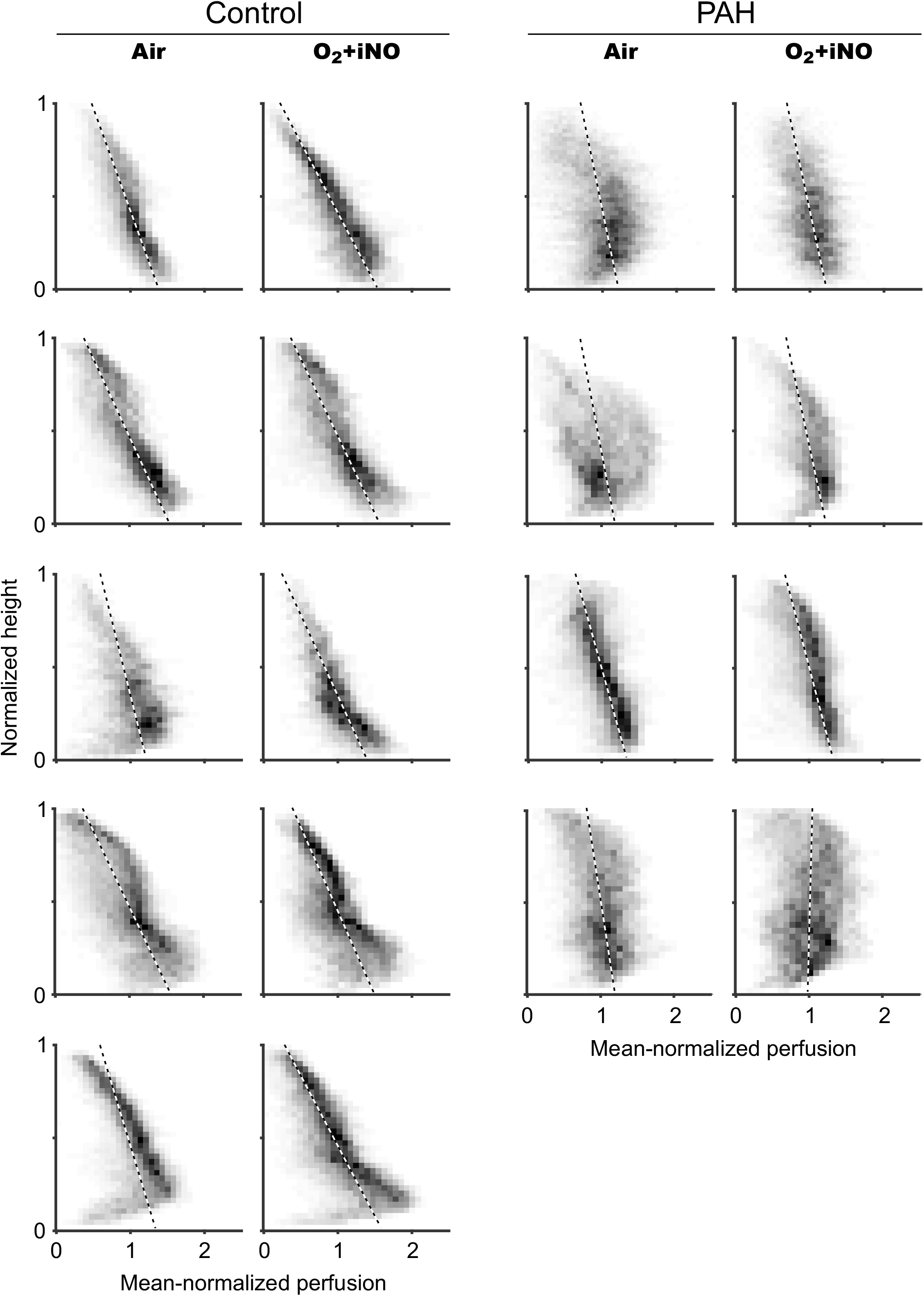
Perfusion-height maps for PAH subjects and controls. The greatest perfusion heterogeneity, including the largest range of perfusion values at any given height, was present in PAH subjects, followed by controls. The vertical gradient in perfusion (represented by the slope of the black-and-white dashed line) increased in magnitude in controls but not PAH subjects breathing O_2_+iNO. In contrast to controls, several PAH subjects had no vertical gradient in perfusion in the lower or dependent part of the lung (dorsal in the supine position), as if the hydrostatic pressure, which increases towards the dependent regions, had no effect on perfusion. Surprisingly, there was no substantial fraction of voxels with very low or zero perfusion in any PAH subject, suggesting there was no severe or complete obstruction of blood vessels. The only confirmed responder of the PAH group is shown in the second row.

The contribution of the axial (craniocaudal) gradient in perfusion to total heterogeneity was small, and there was no significant difference between the groups regardless of O_2_+iNO administration (p=0.17 at baseline, p=0.39 after O_2_+iNO) (Figure 1C).

#### Residual Perfusion Heterogeneity and the Length Scale Spectrum

After removing the vertical and axial gradients in perfusion from the scans, PAH subjects had similar residual heterogeneity (CV^2^_Qr_) when compared to controls (p=0.41) at baseline. These similarities in CV^2^_Qr_ remained consistent while breathing O_2_+iNO. To further analyze the residual spatial heterogeneity in perfusion, we assessed the variations in perfusion across the length scale spectrum. At baseline and while breathing O_2_+iNO, PAH subjects had similar residual heterogeneity across the spectrum of length scales of 10-110 mm (p>0.05 for all included length scales), when compared to control subjects (**Tables 3 & 4**).

**Table 3.** Perfusion Distributions at all Length Scales at Baseline.

| **Parameter** | **Controls** | **PAH** | **p-value** |
| --- | --- | --- | --- |
| **CV<sup>2</sup><sub>Q10-30</sub> (x10<sup>-3</sup>)** | 13.0 (6.82, 29.77) | 19.7 (17.17, 22.25) | 0.33 |
| **CV<sup>2</sup><sub>Q30-50</sub> (x10<sup>-3</sup>)** | 7.77 (4.06, 16.70) | 10.0 (8.05, 11.70) | 0.68 |
| **CV<sup>2</sup><sub>Q50-70</sub> (x10<sup>-3</sup>)** | 4.46 (2.59, 8.15) | 5.61 (4.48, 6.83) | 0.45 |
| **CV<sup>2</sup><sub>Q70-90</sub> (x10<sup>-3</sup>)** | 2.52 (1.69, 3.57) | 3.31 (2.54, 4.67) | 0.20 |
| **CV<sup>2</sup><sub>Q90-110</sub> (x10<sup>-3</sup>)** | 1.48 (0.96, 2.10) | 1.91 (1.41, 3.22) | 0.33 |
| **CV<sup>2</sup><sub>Q>110</sub> (x10<sup>-3</sup>)** | 5.28 (1.19, 16.83) | 5.81 (0.80, 21.04) | 0.80 |

Definition of abbreviations: CV<sup>2</sup><sub>Q(lengthscale)</sub> = squared coefficient of variation in perfusion quantifying
spatial heterogeneity in regional perfusion in the denoted length scale range (mm)

**Table 4.** Perfusion Distributions at all Length Scales at During O_2_+iNO.

| Parameter | Controls | PAH | p-value |
| --- | --- | --- | --- |
| CV<sup>2</sup><sub>Q10-30</sub> (x10<sup>-3</sup>) | 14.4 (6.65, 24.32) | 13.4 (8.24, 17.43) | 0.62 |
| CV<sup>2</sup><sub>Q30-50</sub> (x10<sup>-3</sup>) | 8.43 (4.68, 12.80) | 7.16 (4.66, 9.59) | 0.46 |
| CV<sup>2</sup><sub>Q50-70</sub> (x10<sup>-3</sup>) | 4.62 (2.44, 6.07) | 4.04 (2.50, 6.48) | 0.68 |
| CV<sup>2</sup><sub>Q70-90</sub> (x10<sup>-3</sup>) | 2.61 (1.29, 3.26) | 2.32 (1.48, 3.91) | 0.72 |
| CV<sup>2</sup><sub>Q90-110</sub> (x10<sup>-3</sup>) | 1.51 (0.75, 1.98) | 1.40 (1.04, 2.06) | 0.70 |
| CV<sup>2</sup><sub>Q>110</sub> (x10<sup>-3</sup>) | 9.56 (5.46, 20.68) | 4.66 (4.05, 5.23) | 0.10 |

Definition of abbreviations: CV2Q(length scale) = squared coefficient of variation in perfusion
quantifying spatial heterogeneity in regional perfusion in the denoted length scale range (mm)

#### Change in Perfusion: Results of voxel mapping

PAH and control subjects demonstrated significant changes in regional perfusion between air and O_2_+iNO at the voxel level of the two registered images (**Figures 4 & 5**) without a significant difference between PAH (mean 0.19, range 0.18-0.20) and controls (mean 0.17, range 0.13-0.20). Also, the variation in regional perfusion at the voxel level in each individual subject was higher than the 99.9% confidence interval expected for random differences due to imaging noise, demonstrating that the perfusion distributions during O_2_+iNO at the voxel level were different from those while breathing air.

**Figure 4:**
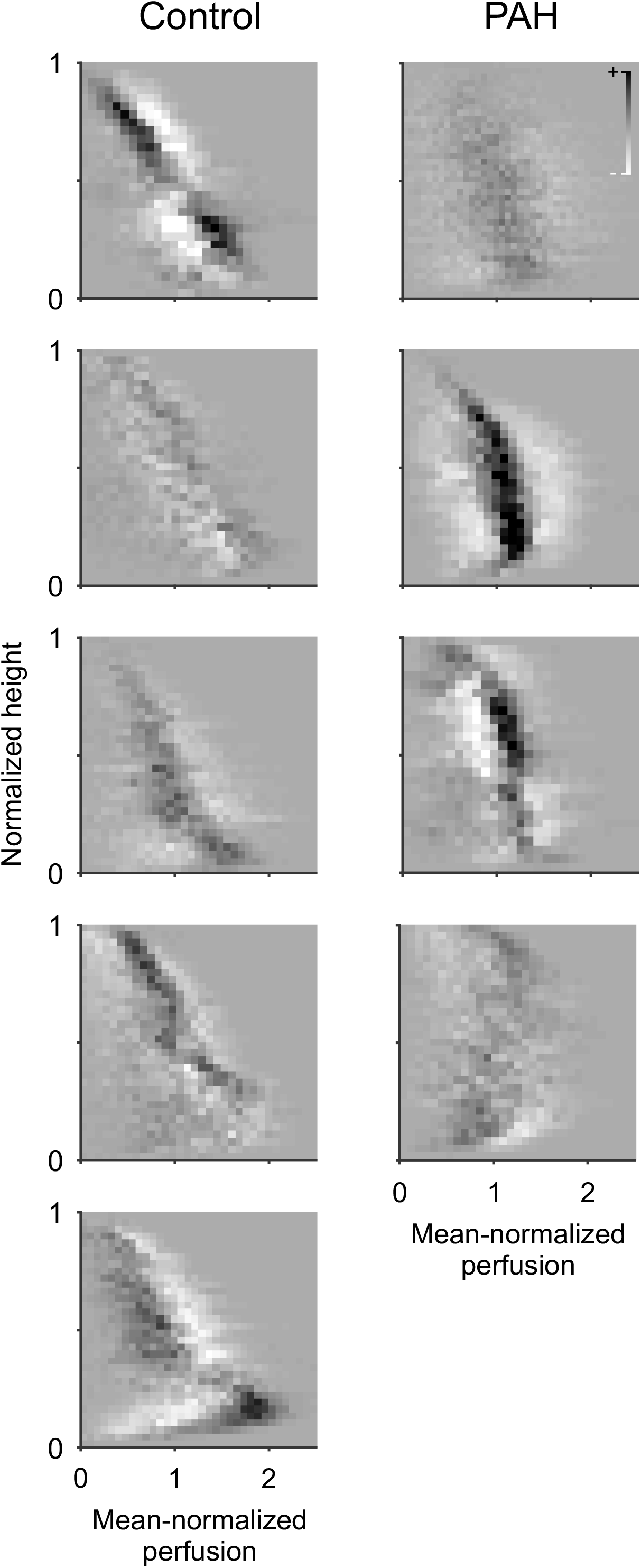
Difference maps of the perfusion-height distribution visualizing the changes in mean normalized perfusion vs. height in response to the administration of O_2_+iNO in PAH subjects and controls. Note that all subjects had regional responses to breathing O2+iNO, including PAH subjects who had no discernible change in mean pulmonary arterial pressures with vasodilator challenge during cardiac catheterization. In controls, the changes in perfusion show a substantial spatial heterogeneity with an overall trend towards an increased vertical gradient during O2+iNO and more uniform perfusion in the dependent (dorsal) regions (see also Fig. 2). In contrast, there are substantial changes in regional perfusion in PAH but not associated with a shift in the vertical gradient except for the confirmed responder among the PAH subjects shown in the second row. All panels use the same scale greyscale for the magnitude of differences.

**Figure 5:**
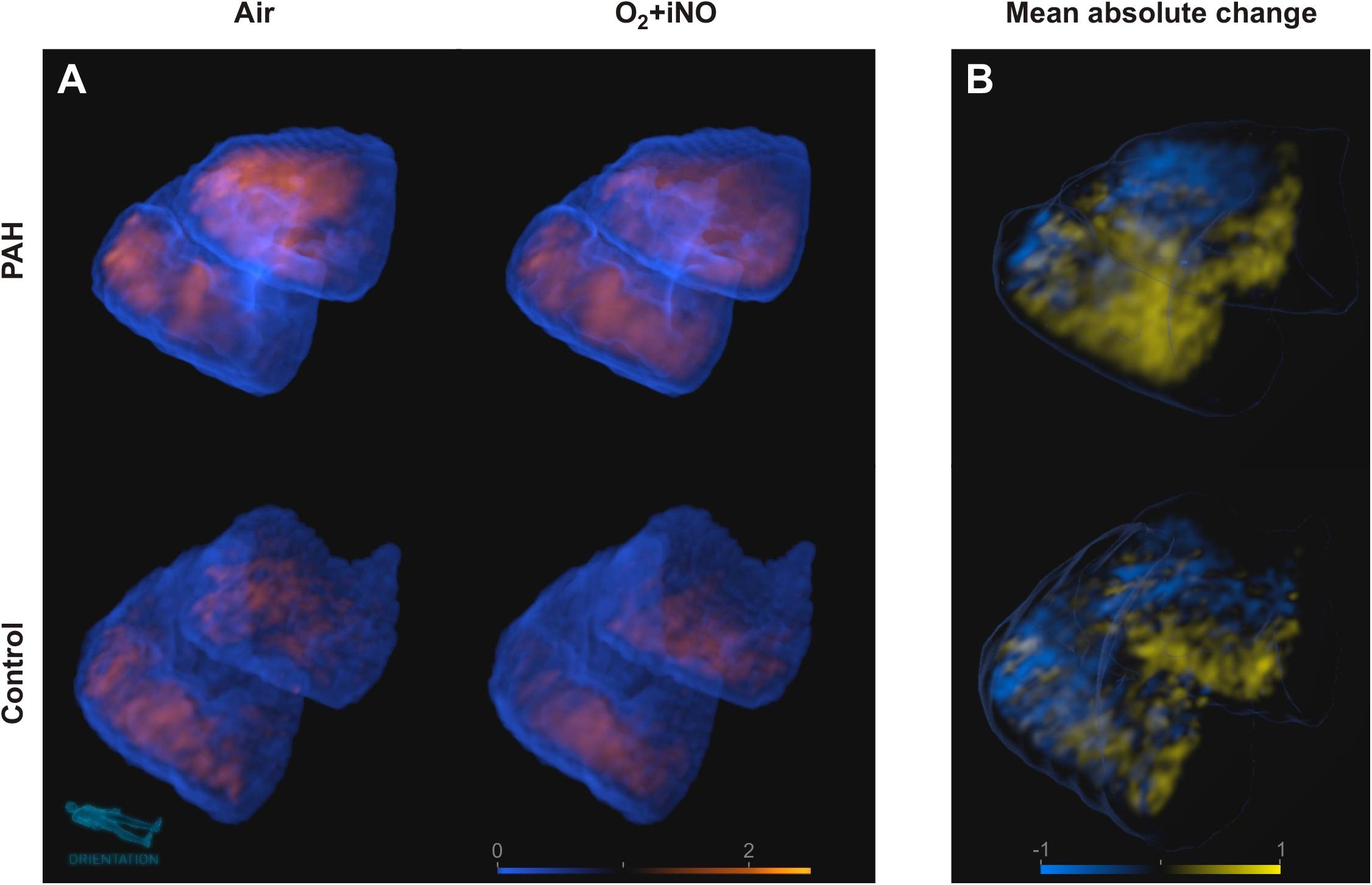
3D renderings and animations of perfusion distributions. **(A)** Representative 3D renderings of the mean-normalized distribution of perfusion for PAH and healthy control subjects at baseline and after O_2_+iNO (animated in Videos V1 (https://doi.org/10.6084/m9.figshare.12442589) and V2 (https://doi.org/10.6084/m9.figshare.12442784)) showing the effects of the administration of O_2_+iNO based on PET-CT imaging in supine position. The patchy areas of high perfusion illustrate the size and location of regional variations behind the representations in perfusion-height maps (Fig. 2). **(B)** 3D renderings of the regional changes in perfusion corresponding to the perfusion distributions in panel A(animated in Videos V3 (https://doi.org/10.6084/m9.figshare.12442787) and V4 (https://doi.org/10.6084/m9.figshare.12442790)). The web-like structure of perfusion increases (yellow) and decreases (blue) instead of gradual changes in perfusion among different regions illustrates the spatial variations behind the difference maps of the perfusion-height distribution (Fig. 3). The relatively small sizes suggest the involvement of smaller anatomical structures together with larger anatomical structures linked to the average changes in larger regions. In all 3D renderings, the orientation of the lungs is shown by a small mannequin. Color scales are fully transparent either for average perfusion (A, V1, and V2) or zero change (B, V3, and V4), opaqueness increases with the deviation from these values, and differences in color between PAH and controls are differences in magnitude. Rotating 3D renderings with periodic switching between air and O2+iNO show the location and changes in high and low perfusion areas. This provides a spatial perception of their location and their magnitude lacking in the still images – video download provides the highest resolution and allows playback in loop mode.

#### Regional Ventilation: Effect of iNO and Oxygen

Alveolar ventilation 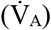 was similar between groups before and during O_2_+iNO (p=0.25, p=0.49 after O_2_+iNO). There was no significant difference observed in the overall mean specific ventilation 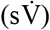 or spatial heterogeneity of 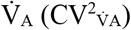 between controls and PAH subjects, reflecting a lack of parenchymal lung disease (**Table 5**). Additionally, lung parenchymal density (estimated by F_tis_) was similar between control and PAH subjects.

**Table 5.** Regional F_tis_ and Ventilation parameters at baseline and during O_2_+iNO.

| Parameter | Control |  | PAH |  |
| --- | --- | --- | --- | --- |
| | Baseline | $O_2+iNO$ | Baseline | $O_2+iNO$ |
| $F_{tis}$ | 0.35 (0.31, 0.43) | 0.39 (0.33, 0.49) | 0.34 (0.27, 0.37) | 0.37 (0.33, 0.41) |
| Volume (ml) | 2107 (1521, 2646) | 1940 (1421, 2347) | 2223 (1676, 3209) | 2147 (1282, 2997) |
| Mean $s\dot{V}$ ($10^{-3}s^{-1}$) | 2.69 (1.49, 4.13) | 2.92 (2.21, 3.51) | 4.68 (1.82, 9.72) | 3.33 (2.00, 5.91) |
| $\dot{V}_A$ (ml/min) | 5.56 (3.73, 8.53) | 5.55 (4.26, 6.72) | 9.36 (4.22, 16.30) | 6.31 (3.85, 7.84) |
| $CV^2_{\dot{V}_A}$ | 0.19 (0.07, 0.33) | 0.18 (0.12, 0.24) | 0.32 (0.07, 0.54) | 0.28 (0.09, 0.37) |

Continuous variables are represented as mean (min, max).
Definitions of abbreviations: PAH = primary pulmonary arterial hypertension; $F_{tis}$ = tissue fraction; Volume = gas volume; $\dot{V}_A$ = alveolar ventilation; $s\dot{V}$ = specific ventilation ( $\dot{V}_A$ /alveolar volume); $CV^2_{\dot{V}_A}$ = squared coefficient of variation quantifying heterogeneity in alveolar ventilation.

#### Ventilation-Perfusion Distributions: Effect of iNO and Oxygen

The distributions of 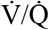 ratios at baseline in PAH subjects were overall broader than in controls at baseline, approaching statistical significance, consistent with moderate degrees of vascular disease-causing mismatch (SD Q 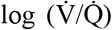, controls 0.51 vs PAH 0.70, p=0.11). There was no difference in the 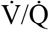 distribution between groups breathing O_2_+iNO, in large part due to widening of the 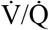 distribution in controls (SD Q 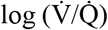, controls 0.65 vs PAH 0.74, p=0.58). The distributions at baseline were unimodal in all subjects, except 2 PAH subjects who had bimodal distributions. 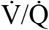 distributions during administration of iNO+O_2_ were unimodal in all subjects including those who had a bimodal distribution prior to the intervention (**Figure 6, Table 5**).

**Figure 6:**
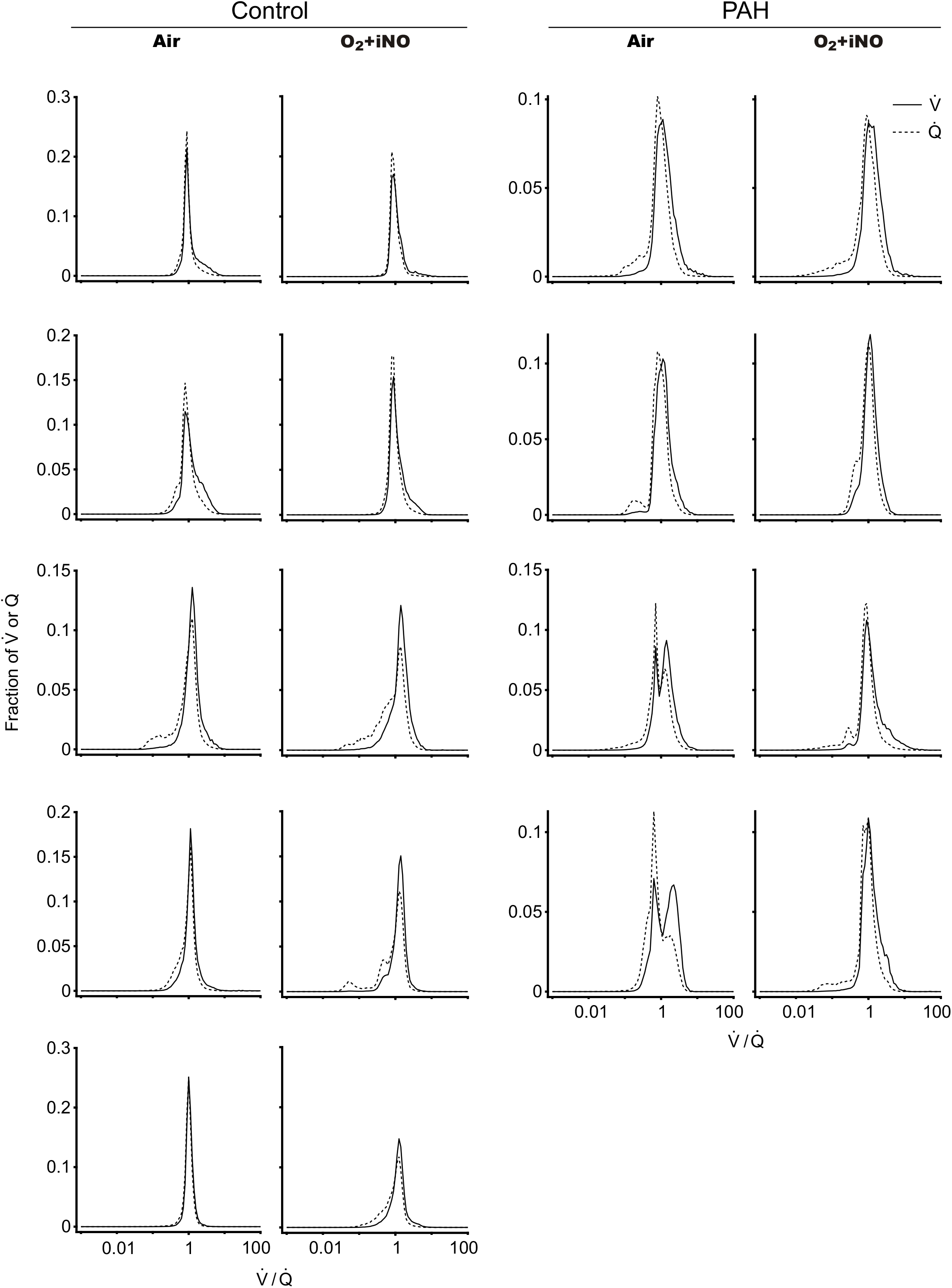
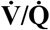 distributions in PAH subjects and controls. The distributions of 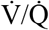 ratios at baseline in PAH subjects were overall broader than in controls at baseline, approaching statistical significance, consistent with moderate degrees of vascular disease, causing a mismatch. There was no difference in the 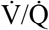 distribution among groups while breathing O_2_+iNO, in large part due to the widening of the 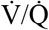 distribution in controls. 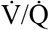 distributions while breathing O_2_+iNO were unimodal in all subjects, including those with a bimodal distribution prior to the intervention.

## DISCUSSION

We report a detailed analysis of regional perfusion and ventilation imaging data in PAH and control subjects obtained with PET-CT, revealing unique perfusion characteristics in PAH subjects that may not be reflected in routine PA pressure measurements. We found that: 1. Breathing O_2_+iNO during imaging resulted in significant differences in the regional distribution of perfusion between PAH subjects and controls, distinguishing the subjects of the two groups. 2. CV^2^_Qvgrad_ during O_2_+iNO had the highest effect size for PAH vs. controls. 3. All subjects in our study responded to O_2_+iNO with changes in regional perfusion, including the two PAH subjects who were non-responders according to vasoreactivity testing. 4. PAH subjects had significantly lower vertical gradients in perfusion than controls and such gradients in 3 of the 4 subjects were unaffected by O_2_+iNO. 5. PAH subjects had residual perfusion heterogeneity across the length-scale spectrum that was similar to that of controls.

An unexpected finding was the magnitude of the vasodilation in healthy controls during O_2_+iNO leading to perfusion redistributions. The significant difference in CV^2^_Qvgrad_ between PAH subjects and controls suggests that perfusion imaging during O_2_+iNO has the potential to be as a biomarker for PAH, that could lead to earlier detection and thus improve patient survival [20]. It remains unclear if the change between O_2_+iNO and air allows identifying responders among PAH patients. The only confirmed responder in our study showed a greater response in CV^2^_Qvgrad_ compared to the other PAH subjects, but it is a single case.

Prior studies using SPECT, MRI and PET have observed that patients with primary precapillary pulmonary hypertension display a pronounced attenuation in the normal gravity-dependent distribution of lung perfusion [13,14,17,19] consistent with our findings. However, these studies reported overlaps between groups, reported only group means, or had not investigated the perfusion distribution during pulmonary vasodilation. One study by Jones et al. [19] using adenosine demonstrated limited change in perfusion redistribution and near absence of a vertical perfusion gradient in PAH even after vasodilation. Based on physiologic principles, studies have hypothesized that since gravity-dependent perfusion redistribution is reliant on low arterial pressures at rest and the ability to recruit and distend vessels with increased cardiac output, higher pulmonary arterial pressures and loss of vascular reserves from obstructive vasculopathies may inhibit vessel recruitment and distension [14,33]. Our study confirmed a lack of significant vertical perfusion redistribution during exposure to O_2_+iNO in those with PAH, indicating that despite measured changes in regional perfusion with vasodilation, PAH subjects did not show normal vertical perfusion patterns. In fact, the vertical gradient in perfusion was in some subjects nearly absent in the dependent part of the lungs, as demonstrated with perfusion-height mapping (**Figure 3**), indicating that vessels at the high pulmonary arterial pressures in PAH may in these subjects already be maximally recruited and distended [34], and therefore are minimally affected by hydrostatic pressures.

An unexpected finding was that the two PAH subjects that were “non-responsive” to vasodilator in terms of the definition of the guidelines for an acute response during vasoreactivity testing [3] had mean values of absolute changes in voxel-by-voxel perfusion during exposure to O_2_+iNO that were similar to those of controls, despite having no change in their vertical gradient in perfusion. Interestingly, there is clinical evidence that patients with a decrease of ≥ 30% in PVR in response to O_2_+iNO benefit from vasodilator therapy although they may not meet the guidelines’ definition for a response during vasoreactivity testing [4]. The hydrostatic pressure gradient caused by gravity would be expected to mainly effect blood vessel distension, depending on the vessel compliance and tone. In this scenario, any loss of vertical gradient could be thought of as a reflection of a loss of overall pulmonary vessel compliance or increase in vessel tone. For instance, any mechanism narrowing the inner lumen, while the outside of the vessel is maximally distended, could explain a high pulmonary vascular resistance, loss of the vertical gradient in perfusion because the hydrostatic pressure gradient cannot expand the vessel, and the lack of a significant change in mPAP to vasodilators. Our data suggests that some degree of vascular tone must be present since we see a change in blood flow during exposure to O_2_+iNO that is similar to controls. Perhaps the increase in tone could be related to the smooth muscle hypertrophy that is seen in PAH [35], and the changes in regional perfusion in our data could explain why some PAH patients can have symptomatic improvement with vasodilators despite little change in mPAP [36].

Our study revealed that PAH subjects had relatively normal values of residual perfusion heterogeneity across the length scale spectrum (**Tables 3 & 4**). This similarity to controls suggests that the vascular disease process may not lead to increased heterogeneity among the resistances of different pathways of blood flow throughout the pulmonary vascular tree. We found the lack of differences in the length scale spectrum very surprising since histopathologic evaluations of the pulmonary vascular tree in patients with PAH have found significant heterogeneity in the type and extent of pulmonary vascular remodelling [37–39] that could have increased the heterogeneity in perfusion at some length scales linked to the anatomical size of the affected sections of the vascular tree, and the length scale spectrum of perfusion heterogeneity has shown differences in other studies, including exercise pulmonary arterial hypertension [12] and COPD [25]. However, histopathologic samples showing structural changes cannot provide complete insights into functional aspects in vivo, such as the vascular resistance along complete pathways through the vessel tree, and systemic consequences of endothelial injury in pulmonary hypertension, such as alterations in the coagulation system, abnormal platelet aggregation and altered production of various endothelial vasoactive mediators, which may be contributing to greater uniformity in the perfusion pattern even in parts of the vascular tree without histopathologic changes. Indeed, although local vascular remodelling and obstruction could be expected to result in patches of low perfusion in PAH subjects, there was no evidence for substantial areas of very low perfusion or significant total spatial heterogeneity of perfusion in our imaging data (Figure 3 & 5). This is relevant because the ^13^NN PET imaging technique measures the regional blood flow through the alveolar capillaries, which is determined by the pressure difference between pulmonary artery and pulmonary vein and the pathway resistance to blood flow. Therefore, the spatial heterogeneity in perfusion includes the overall effects of vascular properties (structural and functional) as well as abnormalities of the vessels along the pathways. In control subjects, O_2_-iNO had no effect on the residual heterogeneity of perfusion CV^2^_Qr_ (Table 2), which is consistent with previous findings that hypoxic pulmonary vasoconstriction had no effect on perfusion heterogeneity in healthy subjects [40].

The relative contribution of gravity and arterial geometry in determining pulmonary perfusion distribution is still subject to debate. Hughes and West argue that gravity contributes to 24-61% of the overall variance in blood flow in the human supine lung [41]. Glenny et al, on the other hand, argue that gravity accounts for a minority of the variability (at most 28%) in perfusion distribution while the geometry of the vascular tree, much of which is genetically predetermined, is claimed to be the primary determinate of pulmonary blood flow [42]. Very few have studied the effects of vasodilation on redistribution of this vertical pulmonary blood flow in healthy subjects. One prior study by Henderson et al. did not find gross redistribution of pulmonary blood flow during hyperoxia in healthy subjects [43] while other studies have found redistribution of blood flow [21] and increased inequality in 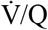 with hyperoxia [44] and iNO [45] in healthy subjects. In our study, vasodilation with O_2_+iNO significantly augmented the vertical gradient of perfusion and widened the 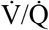 distribution in control subjects. These findings suggest that the controls have baseline pulmonary vasomotor tone enhancing 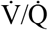 matching which responds to interventions such as hyperoxia and iNO.

The SD Q 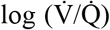 values for controls in our study (0.31-0.81) are consistent with the range of values previously reported in healthy adults using MIGET (0.22-0.72) [46]. Additionally, MRI-based 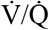 distributions are in good agreement with MIGET measurements [47]. However, MIGET using micropore membrane inlet mass spectrometry (MMIMS) to measure the tracer gases resulted in narrower 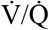 distributions compared to MIGET using gas chromatography [48]. In contrast to MIGET, which involves a smoothing function to estimate the 50 values of a 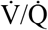 distribution from measurements of six tracer gases, dynamic PET imaging of ^13^NN tracer kinetics allows the estimations of ventilation and perfusion for up to two compartments per voxel and binning of the ventilation and perfusion values to derive 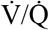 distributions with 100 discrete values without a smoothing function. A validation of the method using heterogenous 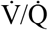 distributions has shown a good agreement between predicted and measured blood gases [49]. In PAH, 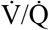 distributions can be substantially wider than we have found [50–53], but our results are consistent with examples of narrower distributions in some PAH subjects [50,51] and the average SD Q 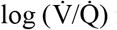 for PAH in our study was slightly higher compared to controls (0.70 vs. 0.51, respectively). We cannot rule out that the exclusion of regions close to the diaphragm where the tidal breathing would cause partial volume effects and axial limitations in the field of view of the PET scanner had some effect on the 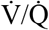 distribution. Additionally, our PET imaging method measures relative pulmonary perfusion and the conversion to blood flow, e.g., in ml/min, would require reliable cardiac output measurements during imaging, which were not available in this study. Our 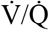 distributions are therefore based on mean normalized ventilation and perfusion.

Despite our study’s strengths, some important limitations should be considered. Limitations of the ^13^NN imaging technique has been discussed in detail [5,27]. Briefly, although PET scans have lower resolution than CT scans, they allow assessment of functional parameters and PET resolution is more than sufficient to quantify functional imaging parameters such as vertical gradients [5]. The effective spatial resolution of the PET perfusion scans is approximately 12 mm, motion blurring from tidal breathing results in lower spatial resolution, but our method of sub-voxel compartments can capture functional heterogeneity at this level. Second, ^13^NN tracer kinetics during the breath hold may affect the accuracy of noise-free CV^2^ parameters of the perfusion heterogeneity, and errors in image registration causing small misalignments could affect the assessment of voxel-by-voxel changes in perfusion. However, such errors only diminish differences among groups and would bias our results toward the null hypothesis of statistical tests. Additionally, an analysis of the reproducibility of CV^2^_Qtotal_, excluding imaging noise, in our previous validation showed a standard deviation of ±8.4% in repeated imaging [28]. Third, despite notable differences observed with this small cohort study, future studies that include other types of patients with pulmonary hypertension and larger studies with matched cohorts will be necessary to assess whether these findings are more generalizable. In contrast to statistical significance, the effect sizes we reported are independent of the sample size. So, larger follow-up studies are expected to confirm our effect size estimates within the reported 95% CIs. A larger cohort should also confirm that the subjects of the small cohort study are representative of the patient population with PAH. Furthermore, additional associations may be present that we are underpowered to detect in this small cohort. For example, it remains unclear if known significant sex differences in cardiovascular parameters [54] and different aspects of PAH [55] are associated with sex differences in regional pulmonary perfusion or not. Similarly, a subgroup analysis comparing responders and non-responders in PAH needs to be conducted in a larger cohort.

## CONCLUSIONS

Perfusion imaging during O_2_-iNO inhalation showed in this small cohort study a large effect size for the heterogeneity associated with the vertical gradient in perfusion, CV^2^_Qvgrad,_ for PAH followed in magnitude by the vertical gradient in perfusion. PAH subjects could be discriminated from healthy controls showing the potential of CV^2^_Qvgrad_ measured during O_2_-iNO inhalation to be a biomarker for PAH. Further studies are needed to investigate NO inhalation in clinically available imaging modalities such as dual-energy CT to translate our findings into clinics.

## Data Availability

The datasets used and/or analysed during the current study are available from the corresponding author on reasonable request.

## ABBREVIATIONS

^13^NN: Isotope nitrogen-13 in a molecule bound to a stable nitrogen atom
2D: two-dimensional
3D: three-dimensional
6MWD: six-minute walk distance
AIC: Akaike Information Criterion
CV^2^: squared coefficient of variation quantifying heterogeneity
CV^2^_Q10-30_: heterogeneity of perfusion at the length scale of 10 to 30 millimeter
CV^2^_Qr_: residual heterogeneity of perfusion
CV^2^_Qtotal_: total spatial heterogeneity of perfusion
CV^2^_Qvgrad_: heterogeneity of perfusion associated with the vertical (dorso-ventral) gradient in perfusion
CV^2^_Qzgrad_: heterogeneity of perfusion associated with the z-axail (cranio-caudal) gradient in perfusion
FEV_1_: forced expiratory volume in first second
F_gas_: gas fraction images using
F_tis_: tissue fraction
FWHM: full-width at half maximum
FVC: forced vital capacity
HRCT: high resolution computed tomography
HU: Hounsfield units
MLV: mean lung volume
mPAP: mean pulmonary arterial pressure
MRI: magnetic resonance imaging
O_2_+iNO: inhalation of nitric oxide (iNO) with balance gas oxygen (O_2_)
PAH: pulmonary arterial hypertension
PCWP: pulmonary capillary wedge pressure
PET-CT: positron-emission tomography and high-resolution computer tomography
PVR: pulmonary vascular resistance
Q_vgrad_: vertical (dorso-ventral) gradient in perfusion
Q_voxel_: perfusion of a voxel
RHC: right heart catheterization
ROI: region of interest
SD Q 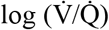: 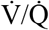 dispersions of perfusion
SD V 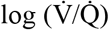: 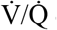 dispersions of ventilation
SPECT: single-photon emission computed tomography
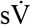: specific ventilation
TLC: total lung capacity
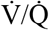: ventilation/perfusion
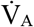: alveolar ventilation

## DECLARATIONS

### Ethics approval and consent to participate

The study was approved by the Institutional Review Board of the Massachusetts General Hospital. Informed consent was obtained from each subject before the study.

### Consent for publication

Not applicable.

## Competing interests

The authors declare that they have no competing interests. For full disclosure, ASW has been consulting for United Therapeutics in the past on topics unrelated to this study, and RC has or had consultant, research, or speaker relationships with United Therapeutics, Janssen, Bayer, Gossamer, Altavant, Third Pole, and Aria CV.

## Funding

Support for this research was provided by an unrestricted grant from United Therapeutics. The sponsor was not involved in the study design, analysis, or reporting of study findings.

## Author information

Tilo Winkler and Puja Kohli contributed equally as joint first authors.

## Contributions

Study design: MK, EGK, RC, RSH; data acquisition: TW, PK, VJK, EGK, JRL, KAH, MK, DMS, JGV, RC, RSH; data analysis: TW, PK, VJK, RSH; data interpretation: TW, PK, RSH; manuscript drafting and editing: PK, TW, VJK, RSH; all authors read and approved the final manuscript.

## Acknowledgements

The authors thank the staff from the Massachusetts General Hospital Departments of Nuclear Medicine and Nuclear Pharmacy for their assistance with this project, including Steve Weise, Michael Cournoyer, John Correia, Eugene Lee, Peter Rice, Melissa Bruen, Erin Beloin, Kevin Vernon and Daniel Yokell. In addition, the authors extend kind thanks to Julian Fischer and Gabriel C. Motta-Ribeiro for their contributions to the process of image registration, Rebecca Harris for her contributions to mask creation, Benno F. Rodemann for his contribution to media creation and to Pradeep Natarajan MD (Massachusetts General Hospital, Division of Cardiology) for helpful discussions and feedback on this work. The present address of Richard Channick is David Geffen School of Medicine at UCLA, Los Angeles, California, USA, and the address of Ekaterina G. Kehl is Division of Pulmonary and Critical Care Medicine, Mount Auburn Hospital, and Harvard Medical School, Boston, Massachusetts, USA.

## Notes

### Competing Interest Statement

The authors have declared no competing interest.

### Summary of Updates

We edited the abstract and the study's limitations to improve transparency and clarity. Additionally, we revised the language of the introduction.

